# The contributions of social factors to lifespan disparity: multi-factor analysis and a four-way case study of U.S. lifespan

**DOI:** 10.64898/2026.03.11.26348159

**Authors:** Hal Caswell

## Abstract

Lifespan varies among individuals, and this variation arises both from heterogeneity (e.g., in sex or race) and from unavoidable individual stochasticity. Treating a heterogeneous population as a mixture of groups, each experiencing different rates, makes it possible to partition lifespan variance into a between-group component due to heterogeneity and a within-group component due to chance. Until now, such studies could only treat factors one at a time. This paper presents an analysis that applies to multi-factor studies and quantifies the contribution of multiple factors and their interactions to variance in lifespan. As a case study it is applied to life table data from a four-factor (sex, marital status, education, and race) study of longevity in the U.S. population, 2015–2019. Even accounting for four main factors, six two-way interactions, four three-way interactions, and one four-way interaction, between-group heterogeneity accounts for only 7% to 10% of lifespan variance. Education and its interactions make the largest contribution. Main effects make the largest contributions; those of two-way, three-way, and four-way interactions are orders of magnitude smaller.

## 1 Introduction

In a homogeneous population, all individuals are subject to the same age schedules of mortality and fertility. Individual lifespan in such a population is a random variable whose probability distribution is given by a column, often denoted *d*_*x*_, of the life table. It is computed from the probability of death, often denoted *q*_*x*_ and the survivorship probability, often denoted by *ℓ*_*x*_. The variability among individuals in this distribution of lifespan is called individual stochasticity [1] or, with some savings of syllables, luck [2].

A heterogeneous population, in contrast, is a mixture of groups, each subject to a different set of demographic rates. Groups may be defined on the basis of measured characteristics (e.g., sex, race, education) or on the basis of latent unmeasured properties (e.g., frailty). Lifespan in such a population follows a mixture distribution in which variability arises from two sources: the differences among the groups and the individual stochasticity that operates within each group.

This variability in lifespan (or any demographic outcome) is will be referred to here as disparity. It is often referred to as an ‘inequality,’ a term that has become politically loaded in some places. Disparity in lifespan has been called the most fundamental of all inequalities because “every other type of inequality is conditional upon being alive.” [3]. Inequality can be quantified in many ways [e.g., 4, Chap. 2], including the variance of the distribution. When measured by the variance, disparity can be partitioned uniquely into two components, one due to differences between groups and one due to variation within groups. These variance partitioning calculations now have a considerable history in studies of longevity and of lifetime reproduction; see Caswell [5] for a review.

Caswell [5] surveys the contributions of heterogeneity (among groups) and individual stochasticity (within groups) to (1) disparity in human longevity among latent frailty classes, income groups, education groups, and neighborhood deprivation groups; (2) disparity in adult longevity among insects differing in early life nutrition; (3) disparity in longevity and lifetime reproduction in a wild seabird population; (4) disparity in longevity and lifetime reproduction in laboratory populations of a rotifer, in which heterogeneity is defined by maternal age; (5) disparity in healthy longevity within and among European countries; and (6) disparity in longevity and lifetime reproduction within and among species of plants and animals. The conclusion from this survey, and of many studies dating back at least to Vaupel [6], is that the contribution of heterogeneity to variance in lifespan is much smaller than that of stochasticity.

However, those studies are limited because they examine only a single source of heterogeneity at a time, although longevity is clearly influenced by many social and biological factors. In this paper, I present an analysis that can incorporate multiple factors and identify the simultaneous contributions of those factors to the variance in any demographic outcome. I apply the analysis to a study of lifespan disparity in the United States from 2015 to 2019, in which heterogeneity in four interacting factors has the opportunity to contribute to the lifespan inequality.

In the next section I present the calculations for multi-factor partitioning of disparity, as a natural extension of the now-classical single factor partitioning. Then the methodology is applied to the data of Bergeron-Boucher et al. [7] on lifespan inequality in the United States. A final discussion section considers at some length some of the issues and questions that arise in partitioning of disparity, and suggests directions for future developments.

## 2 Variance partitioning

Consider a heterogeneous population composed of a mixture of groups, with the representation of the groups given by a probability distribution (the mixing distribution) ***π***. Let *ξ* denote some demographic outcome (here we will be concerned with longevity); the distribution of this outcome is a mixture distribution. The variance in *ξ* is

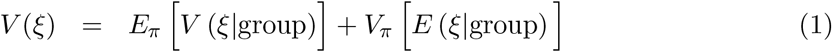

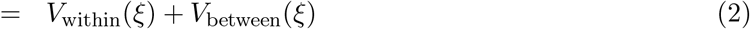

[e.g., 8, 9, Section 5.6]. The within-group variance *V*_within_ is the mean of the variances within each group, weighted by the mixing distribution ***π***. The between-group variance *V*_between_ is the variance of the group means, again weighted by the distribution ***π***. The variance ratio

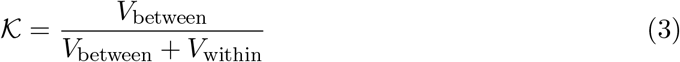

quantifies the proportion of the variance that is due to between-group differences.

The partition (2) has now been applied in many studies and the general conclusion is that for many kinds of heterogeneity the variance ratio is on the order of a few percent, even in the face of heterogeneity in important social factors.

### 2.1 From one factor to many

Studies of lifespan variation have focused on contributions of one factor at a time, using Equation (2) as a tool. For example, Luy et al. [10] reported life tables form men and women as functions of income, education, and occupation, but each factor was treated separately, for men and for women. Caswell [5] used these life tables to partition the variance in longevity into within-group and between-group components, and found that *K* ranged from 0.01 to 0.05 for each of the factors separately.

In a more extensive analysis, van Raalte et al. [11] analyzed lifespan inequality for males and females in eleven European countries and four educational groups, measuring disparity by both Thiel’s entropy-based index and the variance in age at death. Inequality within each of the 88 sex-country-education combinations was analyzed separately. Between-group differences accounted for 0.6% to 11% of the variation in lifespan.

Treating each factor in a multi-factor study design singly rules out the possibility of calculating the contributions to variance of *interactions* between them. The approach presented here, extending that of Caswell and van Daalen [12] makes it possible to partition the variance into contributions from the each factor and from all of their interactions.

### 2.2 Notation

The following notation is used throughout this paper. Matrices and arrays are denoted by upper-case bold characters (e.g., **U**) and vectors are denoted by lower-case bold characters (e.g., **a**). Vectors are column vectors by default; **x**^T^ is the transpose of **x**. The vector **1** is a vector of ones, and the matrix **I** is the identity matrix. When necessary, subscripts are used to denote the size of a vector or matrix; e.g., **I**_*ω*_ is an identity matrix of size *ω* × *ω*. The notation ∥**x**∥ denotes the 1-norm of **x** (i.e., the sum of the absolute values of the entries). The symbol ⊗ denotes the Kronecker product. The vec operator stacks the columns of a *m* × *n* matrix into a *mn* × 1 column vector. When applied to an array with more than 2 dimensions, the vec operator stacks columns from all dimensions. We will make use of a reshape operator (using Matlab notation) that is the inverse of the vec operator, changing the vector back into an array with specified dimensions.^1^

Two element-by-element operations are important in what follows. The symbol ◦ denotes the Hadamard, or element-by-element product (implemented by .* in Matlab and by * in R). The symbol ⊘ is used to denote the less commonly encountered element-by-element quotient. For two matrices **A** and **B**,

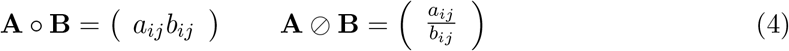

with the obvious restrictions that **A** and **B** must be of the same size and, that for the quotient, none of the entries of **B** can be zero.

### 2.3 Factorial study designs

Consider a study examining the effects of *k* factors, in all possible combinations, on some demographic outcome. In the example to follow, *k* = 4 and the outcome is lifespan. The study includes *n*_1_ levels of factor 1, *n*_2_ levels of factor 2, and so on up to *n*_*k*_ levels of factor *k*.

The means and variances of lifespan, and the mixing distribution, are contained in *k*-dimensional arrays, each of size *n*_1_ × *n*_2_ · · · × *n*_*k*_:

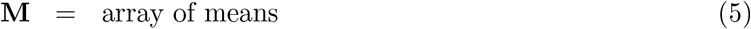

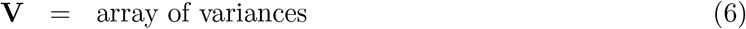

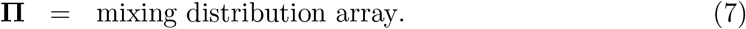

#### 2.3.1 Marginal mean and mixing arrays

The partition of variance is based on the weighted marginal distributions from **M** and **Π**. For notational convenience, let us denote the names of the factors as *A, B, C*, and so on. The entry **M**(*a, b*, …) of **M** is the mean longevity in the group defined by level *a* of factor *A*, level *b* of factor *B*, and so on. Similarly for the variances **V** and the mixing distribution **Π**. We require that

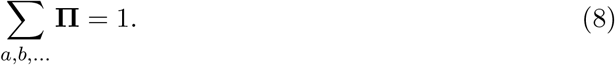

The variance components are calculated from the weighted marginal distributions obtained from the arrays **M, V**, and **Π**. Caswell and van Daalen [12] presented formulas for these marginal distributions for several special cases. To write a general formula, define a set *F* containing all the factors in the study. For example, suppose there are four factors, labelled *A, B, C*, and *D*. Then

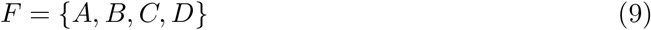

Each possible combination of factors is a partition of the set *F* into a set *S* of factors included in the combination and a complementary set *S*^*c*^ of factors not included. For example the two-way interaction of A and C is defined by

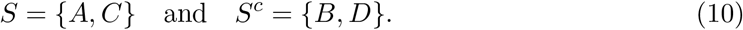

Using these sets, the marginal mean array and the marginal mixing distribution array for the interaction among the factors in the set *S* are

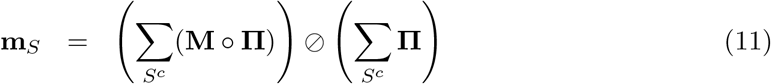

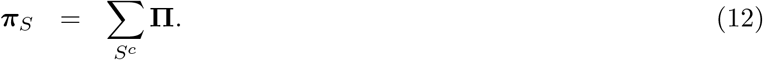

where ⊘ is the Hadamard, or element-by-element quotient. For example, the marginal *AC* mean and mixing distribution, with *S* and *S*^*c*^ given by (10) are

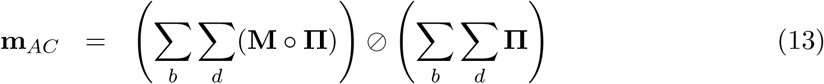

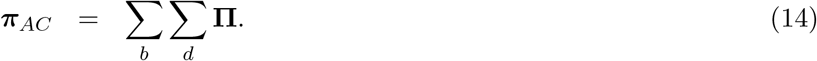

#### 2.3.2 Within-group and between-group variances

The study design contains *n*_1_*n*_2_ · · · *n*_*k*_ factor combinations. By treating each of these combinations as a group, the analysis is reduced to a one-factor design. Then, as in Equation (2), the within-group variance is given by the expectation, over the mixing distribution, of the variances in the groups:

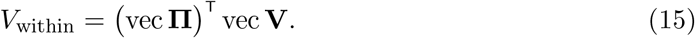

The between-group variance is the variance, over the mixing distribution, of the group means. In general, we can define the variance of the entries of a vector **x** over a distribution ***π*** as the function

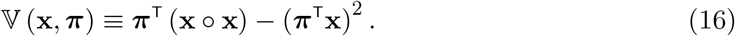

The first term in V (·, ·) is the second moment (the expectation of the squares of the entries of **x**). The second term is the square of the first moment. In terms of this function, the overall between-group variance is

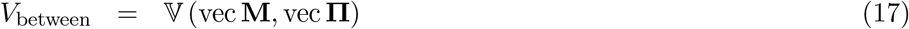

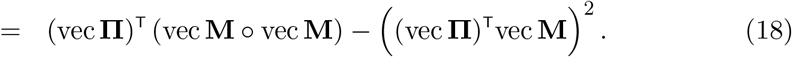

#### 2.3.3 Partitioning variance: main effects and interactions

In a study including *k* factors, the between-group variance *V*_between_ in Equation (17) can be partitioned into *k* single-factor contributions, 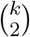 two-way interactions, 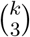 three-way interactions, and so on up to a single *k*-way interaction.

The variance contribution of each of these interactions is defined in terms of the set *S* as

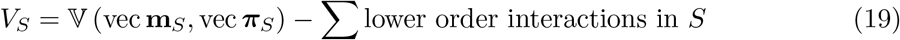

where **m**_*S*_ and ***π***_*s*_ are the marginal mean array and marginal mixing array corresponding to the set *S*, calculated according to (11) and (12).

The case study to be examined here contains four factors: sex, marital status, education, and race (see Section 3), labelled A, B, C, and D. The between-group variance in lifespan is partitioned into components due to four single-factor effects (*V*_A_, *V*_B_, *V*_C_, *V*_D_), six two-way interactions (*V*_AB_, *V*_AC_, *V*_AD_, *V*_BC_, *V*_BD_, *V*_CD_) between pairs of factors, four three-way interactions (*V*_ABC_, *V*_ABD_, *V*_CD_, *V*_BCD_) and one four-way interaction (*V*_ABCD_).

Examining a single example of each order of interaction reveals the general pattern. The explicit formulas for all the variances are given in Appendix **??**.

- Single-factor variances (e.g., *V*_A_)

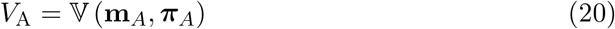
- Two-way interactions (e.g. *V*_AB_)

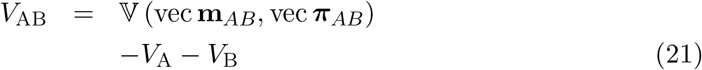
- Three-way interactions (e.g., *V*_ABC_)

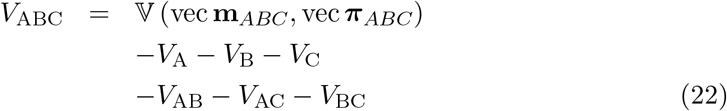
- Four-way interaction (e.g., *V*_ABCD_)

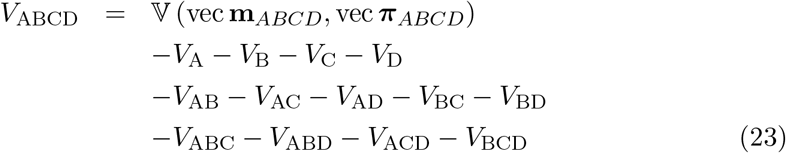

In each case the variance calculated from the marginal distributions by the function V(·, ·) includes the effects of all lower-order interactions, and so those must be subtracted from V (·, ·) to obtain the variance *V*_*S*_.

### 2.4 Choosing the mixing distribution

As discussed in Caswell and van Daalen [12] (and as familiar from experimental design texts), it is only possible to partition variance into its components if the mixing distribution is either uniform (or “flat” in the usage here) or a rank-one, proportional distribution. A flat mixing distribution gives equal weight to every factor combination. In a rank-one distribution, each column of **Π** is proportional to every other column and each row is proportional to every other row. Thus the flat distribution is a special case of a rank-one distribution.

A rank-one mixing distribution is assembled from its marginals as follows. Consider the case with four factors, and let ***π***_*A*_, ***π***_*B*_, ***π***_*C*_, and ***π***_*D*_ be the marginal distributions over levels of *A, B, C*, and *D* respectively. The rank one mixing distribution array with these marginal is

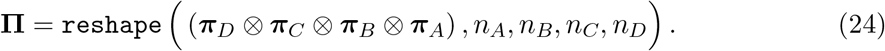

The Kronecker product term creates a vector of dimension *n*_*A*_*n*_*B*_*n*_*C*_*n*_*D*_ × 1. The `reshape` operator assembles the entries of this vector into a 4-dimensional array of dimension *n*_*A*_ × *n*_*B*_ × *n*_*C*_ × *n*_*D*_. Note the order of the terms.

Flat and rank-one mixing distributions ask, and answer, different questions. Neither is right or wrong. For more discussion, see Section 4.1.

## 3 A case study: lifespan inequality in the United States

Bergeron-Boucher et al. [7] presented an analysis of lifespan and mortality risk in the United States from 2015 to 2019. Their study is unique because they reported life tables for all 54 combinations of four factors:

| Factor | Levels |
| --- | --- |
| A: sex | female, male |
| B: marital status | married, previously married, never married |
| C: education | high school degree or less, some college, college degree |
| D: race | Black, Hispanic White, non-Hispanic White |

The life tables were constructed starting at age 30 and were truncated at age 90. They were based on data from the Multiple Causes of Death Dataset of the National Center for Health Statistics and the American Community Survey. Data were pooled from 2015 to 2019; years from 2020 onward were excluded to avoid effects of the COVID-19 pandemic. For further details see Bergeron-Boucher et al. [7]. They analyzed partial life expectancy between ages 30 and 90, seeking to identify factors influencing that index, and related it to causes of death. They report the population sizes of each of the 54 groups, which we have used to construct one of the weighting distributions.

These four factors are of undoubted social and demographic significance. Their combinations make a sizable impact on life expectancy: Bergeron-Boucher et al. [7] report an 18-year difference between the factor combination yielding the lowest and the combination yielding the highest partial life expectancy, a result that is replicated in our calculations.

These data provide an ideal opportunity to explore lifespan disparity by partitioning the variance in longevity into contributions from each factor and from the six two-way, four three-way, and one four-way interaction among the factors. For the analysis here, means and variances in remaining longevity were computed from each life table using Markov chain methods. These means and variances were used in constructing the arrays **M** and **V** in equations (5) and (6).

The variance components for all main factor effects and all interactions were computed, as in Section 2.3.3, using both the flat and a population-weighted mixing distribution. In the flat distribution **Π**(*a, b, c, d*) = 1*/*54. The population-weighted distribution was assembled as a rank-one distribution from the marginal population sizes over each factor, using Equation (24). Figure 1 displays these mixing distributions graphically, emphasizing the weight given to the various factor combinations, from the largest group to the smallest.

**Figure 1.**
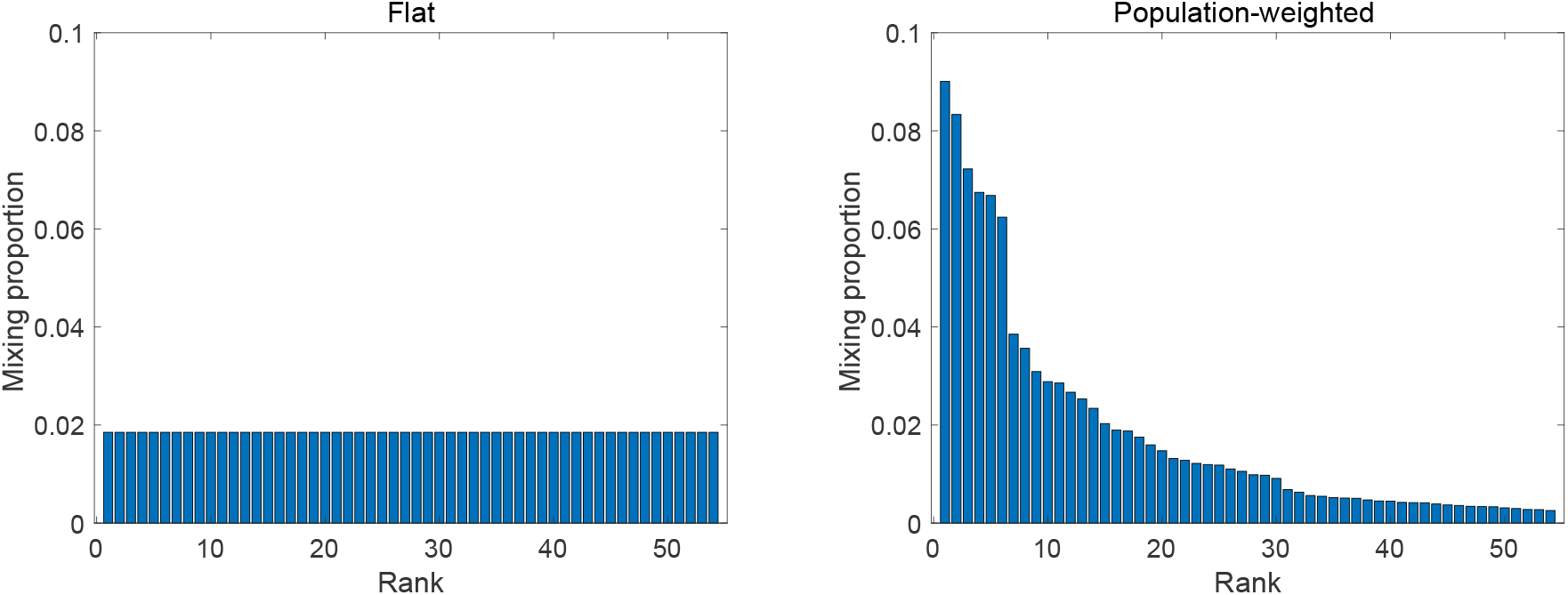
The representation of the 54 factor combinations, ranked from highest to lowest, in the flat and the population-weighted mixing distributions.

### 3.1 Components of variance

The variance components can be presented in various ways. I present several of these here because they highlight different aspects of the same results. A complete tabulation of the variance contributions from all main effects and interactions, for both mixing distributions, is given in Table 1. The between-group variance *V*_between_ contributes only 11% (flat) or 8% (pop-weighted) of the total variance. The remainder is due to individual stochasticity.

**Table 1.** Components of variance in remaining longevity due to sex, marital status, education, and race, and the percent of the total variance contributed by each. Starting age = 30. Results are shown for flat and for population-weighted mixing distributions.

| Component | Flat mix |  | Pop. mix |  |
| --- | --- | --- | --- | --- |
|  | Variance | Percent | Variance | Percent |
| A=sex | 3.94 | 2.2 | 2.64 | 1.8 |
| B=marital status | 3.96 | 2.2 | 2.95 | 2.0 |
| C=education | 8.81 | 4.9 | 4.78 | 3.2 |
| D=race | 0.71 | 0.4 | 0.70 | 0.5 |
| AB=sex × marital status | 0.12 | 0.07 | 0.09 | 0.06 |
| AC=sex × education | 0.31 | 0.17 | 0.20 | 0.13 |
| AD=sex × race | 0.03 | 0.02 | 0.01 | 0.01 |
| BC=marital status × education | 0.34 | 0.18 | 0.12 | 0.08 |
| BD=marital status × race | 0.37 | 0.20 | 0.13 | 0.09 |
| CD=education × race | 0.35 | 0.20 | 0.12 | 0.08 |
| ABC=sex × marital status × education | 0.02 | 0.01 | 0.01 | 0.01 |
| ABD=sex × marital status × race | 0.04 | 0.02 | 0.04 | 0.02 |
| ACD=sex × education × race | 0.03 | 0.02 | 0.01 | 0.01 |
| BCD=marital status × education × race | 0.10 | 0.05 | 0.06 | 0.04 |
| ABCD=sex × marital status × education × race | 0.01 | 0.01 | 0.00 | 0.00 |
| $V_{\text{between}}$ | 19.15 | | 11.86 | |
| $V_{\text{within}}$ (stochasticity) | 161.25 | | 138.45 | |
| Total | 180.39 |  | 150.72 |  |
| $\mathcal{K}$ | 0.11 | | 0.08 | |

Education makes the largest contribution, accounting for 4.9% (flat) or 3.2% (pop-weighted) of the total variance. Sex and marital status contribute similar amounts, with race a distant third. The contributions of interactions are smaller than the contributions of the main effects.

Interactions are of particular interest, because they draw attention to the ways in which the response to one factor depends on the value of a different factor. Table 2 displays the contributions of the main effects and interactions, grouped by the order of the effects. The contributions of the two-way interactions are an order of magnitude smaller than those of the main effects. The three-way interaction contributions are an order of magnitude smaller yet.

**Table 2.** Components of variance in remaining longevity due to sex, marital status, education, and race. Main effects and interactions, grouped by two-way, three-way, and four-way. Starting age = 30 years. Results are shown for flat and for population-weighted mixing distributions.

| Component | Flat mix | Pop. mix |
| --- | --- | --- |
|  | Variance | Variance |
| Main effects | 17.42 | 11.07 |
| Two-way interactions | 1.52 | 0.67 |
| Three-way interactions | 0.19 | 0.11 |
| Four-way interaction | 0.01 | 0.00 |
| $V_{\text{between}}$ | 19.15 | 11.86 |
| $V_{\text{within}}$ (stochasticity) | 161.24 | 138.45 |
| Total | 180.39 | 150.72 |
| $\mathcal{K}$ | 0.11 | 0.08 |

The contributions of all factors and interactions are smaller under the population-weighted mixing distribution than under the flat mixing distribution. This reflects the greater concentration of the population-weighted mixing distribution into the most abundant of the 54 groups (Figure 1.

Even when, as here, heterogeneity creates large differences in survival and life expectancy, the contribution of that heterogeneity to lifespan disparity is small, being overwhelmed by individual stochasticity. It may be useful to focus on the importance of the factors as measured by their contributions to the between-group variance *V*_between_ rather than the total variance. Table 3 shows these contributions. The main effect of education contributes 46% (flat) or 40% (pop-weighted) of the between-group variance. Sex and marital status each contribution about 20% and race contributes 3.7% (flat) or 5.9% (pop-weighted).

**Table 3.**
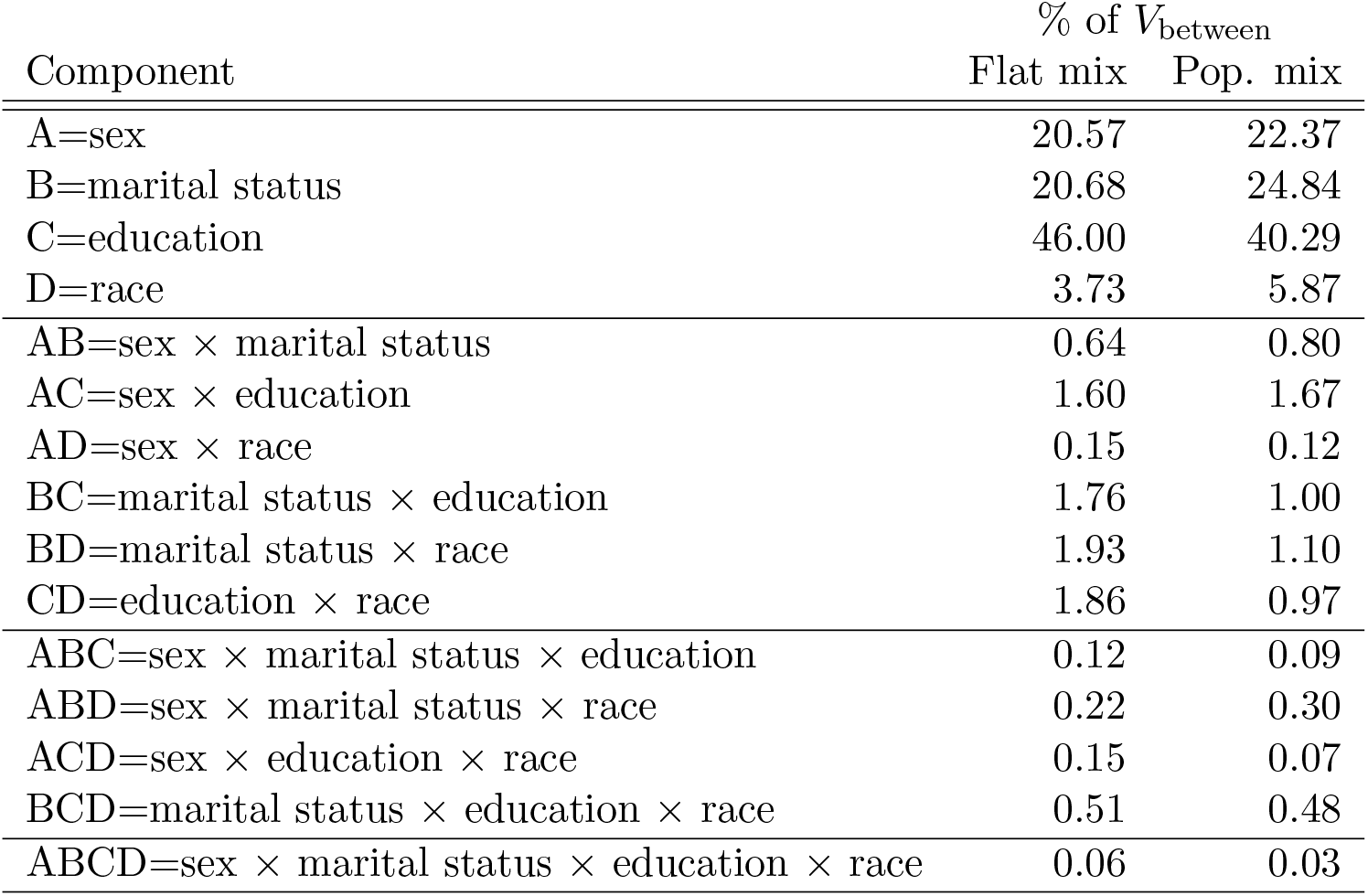
Components of variance in longevity due to sex, marital status, education, and race. Variance components expressed as percentages of *V*_between_. Results are shown for flat and for population-weighted mixing distributions.

### 3.2 Importance of factors: Sobol’ global sensitivity

The importance of education as a factor can be formalized using the Sobol’ global sensitivity index [13, 14]. If some quantity is a function of a set of variables *x*_1_, *x*_2_, …, Sobol’ argued for using the components of variance as a measure of the importance of the various factors [15]. The Sobol’ index S of a factor is obtained by summing the variances due to the main effect of the factor and all the interactions that include that factor. In our case

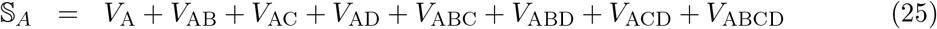

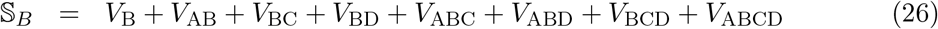

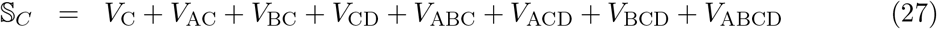

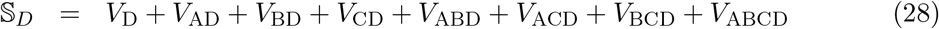

Table 4 shows the Sobol indices for each of the four factors. Under either mixing distribution the Sobol’ index for education is the largest. This supports other claims about the importance of education in determining longevity [e.g., 16, 17, 11]. The indices for sex and marital status are smaller and similar to each other, showing the two factors as similarly important in determining longevity.

**Table 4.**
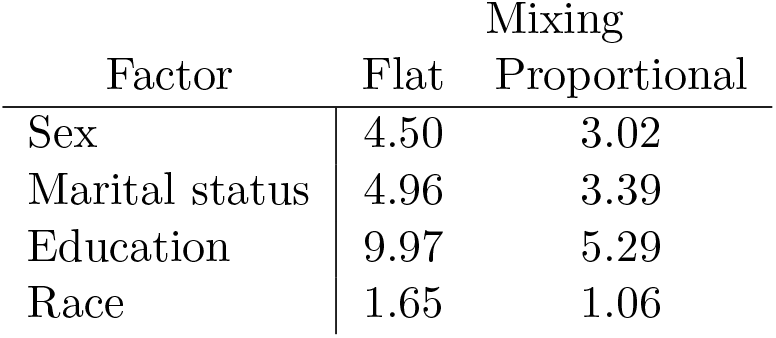
Sobol’ indices for the four factors. The index gives the contribution to variance of the factor, pooling main effects and all interactions involving that factor. Initial age = 30.

### 3.3 Inequality as a function of age

The variance in remaining lifetime can be calculated conditional on any starting age. Figure 2 shows the within- and between-group contributions, *V*_within_ and *V*_between_, as a function of age. Under either mixing distribution, the total variance and the between-group variance decline with age. The between-group component never becomes larger than at age 30, which is the first age reported in the data.

**Figure 2.**
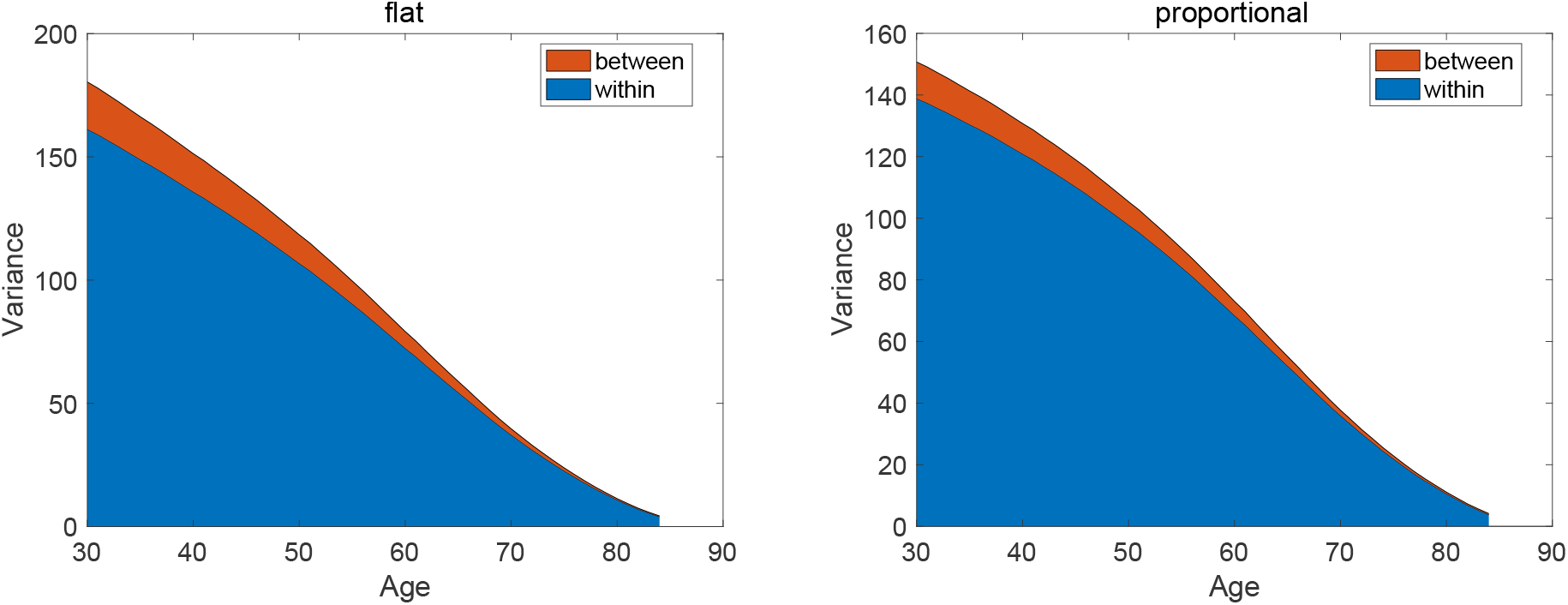
Within-group *V*_within_ and between-group *V*_between_ variances in remaining longevity, as a function of age, for flat and population-weighted mixing distributions.

The variance ratio *K* declines with the starting age of the calculation up to about 80 years, after which it increases (Figure 3) . The final increase may reflect the truncation of the life tables at age 90, because at ages approaching this maximum *V*_within_ must decline towards zero.

**Figure 3.**
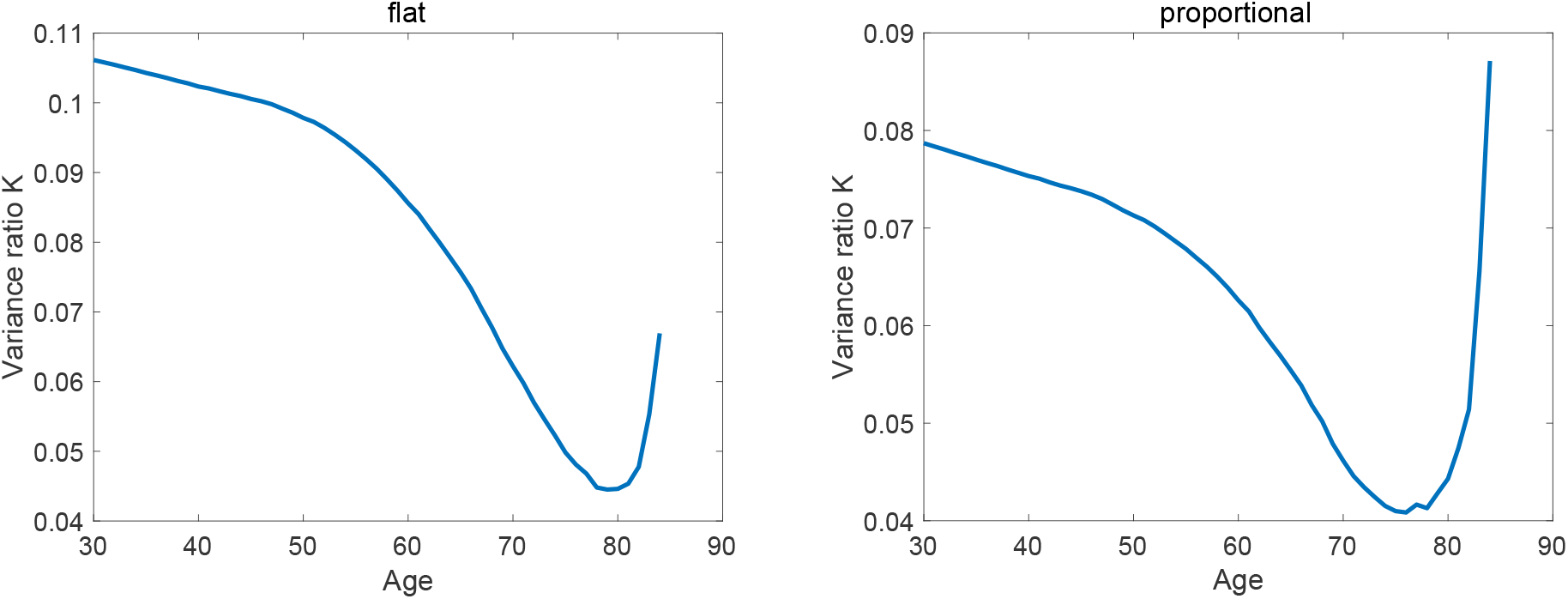
The variance ratio *K* for remaining longevity as a function of starting age, for flat and proportional mixing distributions.

## 4 Discussion

The ability to partition variance in multi-factor studies opens new perspectives on the contributions of social and biological factors to lifespan disparity, and how that is related to inequalities in those factors. As shown in Section 2.3.1, the components of variance due to each factor and to all of their interactions can be written in terms of the partitions of the set of factors and a function giving the variance of the entries of a vector. The terms of this partition account for the simultaneous operation of all the factors, rather than treating each one separately.

In the present four-way factorial case study, the contributions of heterogeneity to lifespan variance are typical of those found in single-factor studies. Note that this is a novel empirical finding; it need not have been true. Even with four factors and 11 interactions, the variance ratio *K* is still small, on the order of 8–10%. About 90% or more of the variance in remaining longevity is due to stochasticity. The contributions to inequality of the factor interactions are even smaller. This is also a novel empirical result. It is not difficult to imagine interactions among sex, marital status, race, and education that might have made a large contribution to variance, but this did not happen.

Within-group variance due to stochasticity is large and, because lifespan is the result of a repeated random outcome of survival, unavoidable. This suggests that it may be interesting to evaluate factors not by their contribution to total variance, but by their contribution to the between-group variance, thus removing stochasticity from the picture. In such a comparison (Table 3), education and interactions involving education make the biggest contributions, contributing 52% of the between-group variance under a flat mixing distribution and 44% under a population-weighted mixing distribution.

### 4.1 Mixing distributions

Mixture distributions are a powerful tool for statistical analysis of heterogeneous populations; see Everitt and Hand [18] or Frühwirth-Schnatter [8] for surveys. The mixing distribution ***π*** is an indispensable component of the description of a heterogeneous population. Although mixing distributions are, in general, enormously flexible, the goal of partitioning variance in multi-factor studies imposes the restriction to flat or rank-one distributions.

The idea that one can choose among multiple possibilities for a mixing distribution deserves clarification. In a flat mixing distribution, each factor combination receives the same weight in the calculation of means and variances. This maximizes the information obtained about the effects of the factors involved. Imagine that you wanted to know how sex, marital status, education, and race, as factors, contribute to inequality in longevity. Imagine that you collect 54,000 individuals and randomly assign 1000 to each of the 54 combinations of those factors.^2^ This flat mixture over the factor combinations would maximize the information about the contributions of each factor provided by the study.

Alternatively, you might sample 54,000 individuals from a population, and assemble the mixing distribution from the relative abundance of the factor combinations in that sample to assemble the mixing distribution. In this calculation, some groups would have more influence than others because they are more common in the population. In the case study analyzed here, the largest group (married white males with a university degree) is 82 times larger than the smallest group (previously married Hispanic males with a university degree). If your goal is to draw conclusions about the sources of inequality in a population that has this composition, then the population-weighted mixing distribution is your preferred tool.

Branches of science in which experiments like the imaginary one described above are regular occurrences have long had to confront this issue. Kendall and Stuart [19, pp. 25–26] describe the choices particularly clearly.

> “In many experimental contexts there is no question of the observations being a random sample from some population. The *r* × *c* cross-classification is deliberately set up to throw light on the variable (*y*) being studied. It may even be meaningless to consider any set of weights as the ‘right’ ones, in the sense of reflecting an underlying population distribution; for example, if we have a 2 × 3 cross-classification to study the effects of two different doses of Fertilizer A and three different doses of Fertilizer B on the yield of a crop (*y*), one may be simply interested in the effects and interactions as such, and not as representing any population at all.
>
> “There is a crucial distinction here between the “experimental” and the “survey” approach to data … In experimental investigations, therefore, it is common (for lack of any known appropriate system of weights) to use equal weights throughout. The distinction between these two weighting systems will become acute, if only because, in this most general case, we may have a very small number of very large frequencies which tend to dominate the frequency weighting system, and perhaps distort its interpretation.”

Kendall and Stuart‘s example of two types of fertilizer in an agricultural experiment corresponds to our position if we view sex, marital status, education, and race as factors and want to evaluate their effects on lifespan variation. This leads naturally to the flat mixing distribution. Evaluating these effects is a different question from evaluating the role of the factors conditional on the composition of the U.S. population. Treating the results as a sample from that population leads naturally to a mixing distribution weighted by population size. The distortion due to a small number of large frequencies mentioned by Kendall and Stuart corresponds to that due to the 82-fold difference between the most and least abundant groups in this analysis.

It is possible to create other rank-one mixing distributions, assembled from marginal distributions as in Equation (24), based on criteria other than population size. The use of such distributions is an open research question.

In the end, the important message is that in analyzing contributions of heterogeneity and stochasticity, the mixing distribution is an analytical tool that we can deploy to answer more than a single question.

### 4.2 Stochasticity is unavoidable; we should embrace it

> *My apologies to chance for calling it necessity*,
>
> *My apologies to necessity if I’m mistaken, after all*.
>
> Wislawa Szymborska, *Under One Small Star*

The life table is a stochastic model for individual mortality. The variance in longevity within any homogeneous population is due to the individual stochasticity inherent in the life table, and is calculated as part of basic survival analysis. It is tempting to imagine that some invisible hidden heterogeneity could contribute to this within-group variance, and explain it as due to something other than chance. But this is impossible until that source is included as a factor, for any analysis based on *calculation* of means and variances of longevity from a life table or a Markov chain.

The logic is different for empirical measurement, rather than calculation, of the distribution of longevity [5]. The differences among individuals in an observed frequency distribution of age at death arise both from stochasticity and from whatever heterogeneity exists among those individuals. Estimation of latent heterogeneity (e.g., frailty; see [20]) make it possible to partition some of that unobserved heterogeneity [21].

Stochasticity produces inequality of outcome. Heterogeneity produces inequality of opportunity, because individuals in different groups experience lives governed by different sets of rates. How to respond to these types of inequalities, especially those due to chance, is a non-trivial problem [5, 22]. Frank [23] surveys arguments about the relative importance of effort and talent on one hand, and luck on the other, in determining individual economic success. Conservative positions emphasize the former, liberal positions the latter. Behavioral experiments show that there is a tendency to underestimate the role of chance in success. The large contributions of stochasticity to inequality of lifespan does not mean that social, behavioral, and biological factors are not worthy of attention.

### 4.3 What if we have individual measurements?

Although the partition of variance results in Table 1 is reminiscent of the results of statistical Analysis of Variance (ANOVA), the method used to get those results is fundamentally different. As is well known, ANOVA begins with a linear model,

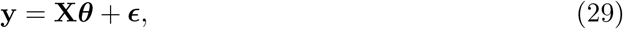

where **y** is a vector of observations, ***θ*** is an unknown vector of parameters **X** is a known design matrix, and ***ϵ*** is a vector of random deviations. Because there are more observations than there are parameters to be estimated), there is no solution to the equation an estimate 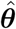 is found by minimizing the sum of squared deviations between observations and estimates. The vector **y** contains individual observations. In our context these would be observations of individual lifespans, but the life tables that form the basis of much of demography contain no such individual measurements. Instead, life tables are computed from counts of deaths and of individuals at risk of death at each age. Treating the moments of longevity as describing a mixture distribution permits the variance decomposition without individual observations.

However, with considerable research effort it is possible to extract individual longevity data from published records; this has been done in the CenSoc project at the University of California at Berkeley [24]. That project links U.S. census and Social Security records for the cohort of men born in 1910 and tracks them until their deaths between ages 65 and 95. The Census data provide information on a variety of social and environmental variables on these individuals. Breen and Seltzer [25] use these data to attempt to predict individual lifespan on the basis of a set of 32 variables (including detailed information on education, race, occupation, household structure, and movements). Rather than a simple linear model, they deployed an ensemble of machine learning algorithms to find a function predicting individual mortality from these variables (full details in their paper). They found statistically significant relations between the variables and individual lifespan but that relationship explained only 1.3% of the variance. As with the case study reported here, they found education-related factors to be the most important, but those explained only 0.8% of the variance in age at death.

Although detailed individual data sets (census records, registers) will yield increasingly detailed predictions of individual mortality, it seems unlikely that simply knowing more and more about everybody will eliminate the importance of individual stochasticity.

### 4.4 Measures of inequality

Economists have employed a great variety of measures of economic inequality [4]. These have been applied in various demographic contexts; when applied to a single population, the results are highly correlated [e.g., 26, 27, 28]

But comparing two or more groups in a heterogeneous population requires a measure that can be partitioned into contributions that reflect that heterogeneity, being sensitive to inequality both within and between the groups. Of the indices in common use, the variance and entropy are the only indices that permit this partition.^3^ The standard deviation is sometimes used on the grounds that it is measured in the same units as the outcome (years, in the case of longevity). However, it cannot be partitioned. The standardized variance, defined as the ratio of the variance to the square of the mean, has been used, and it can be partitioned [29], making it preferable to the coefficient of variation, which is its square root. Its use in multi-factor study designs is still to be explored. The Gini coefficient, which is based on the mean absolute difference among individuals, is popular for studies in a single population. However, it cannot be partitioned into within-group and between-group components.

### 4.5 Possible extensions

Variance partitioning can be applied to any demographic outcome for which means and variances can be calculated. In addition to longevity, other obvious candidates include healthy longevity [30], state occupancy times in multistate models [31], lifetime reproductive output [32, 2, 33], and the abundance of, and lifetime overlap with, various types of kin [34, 35].

Extending the mixing distribution beyond the flat and the population-weighted cases provides a novel opportunity to address longevity from other perspectives. The case study here uses a weighting distribution proportional to the population sizes reported by Bergeron-Boucher et al. [7] for each of the 54 groups. Other quantities could be used. Population size of a particular age classes an obvious choice. Mixing distributions might also be defined based on the number or proportion of individuals with particular religious or political affiliations, or levels of wealth, income, or debt. Exploring these is an open research area.

Factorial study designs are those that include all possible combinations of the factors studied. Other multi-factor designs are possible, in particular hierarchical designs in which groups are composed of sub-groups, each of which is composed of sub-sub-groups, and so on.The largest demographic data sets in the world (e.g., the U.N. World Population Prospects) have such a hierarchical structure, in which geographical regions are composed of subregions which in turn are composed of countries. Variance partitioning in such studies are also possible and will be presented elsewhere (Caswell, unpublished results).

## Data Availability

All data produced in the present work are contained in the manuscript.

## Ethical review

This study uses only anonymous statistical summary data which include no individually identifiable elements. Hence, ethical review board approval is not needed.

## Informed consent

Not applicable.

## 5 Acknowledgements

I thank Marie-Pier Bergeron-Boucher for generously providing the life table data used in these calculations. I am happy to acknowledge the late Dr. John L. Gill of Michigan State University.

## Appendix

### A Complete formulas for four-way variance components

The four factors are labelled *A, B, C*, and *D*. The between-group variance is partitioned into four single factor main effects, six two-way interactions, four three-way interactions, and one four-way interaction.

- Single factor contributions:

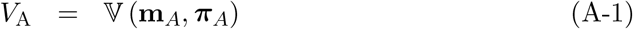

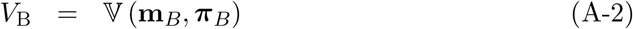

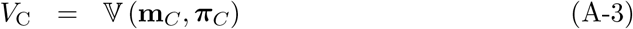

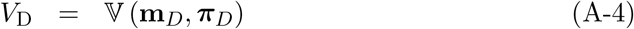
- Two-way interactions:

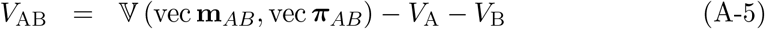

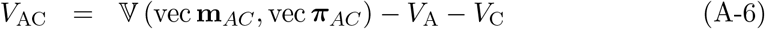

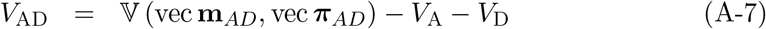

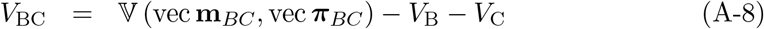

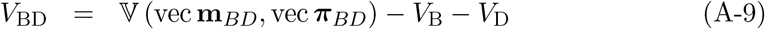

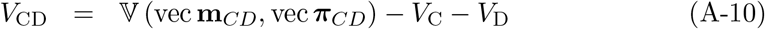
- Three-way interactions:

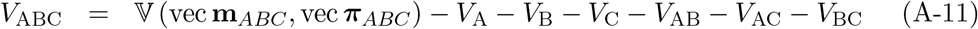

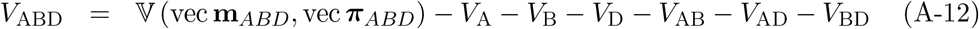

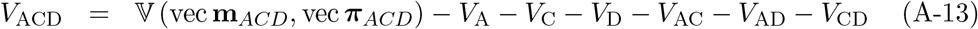

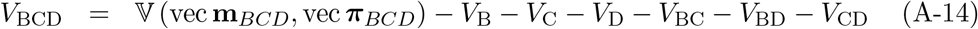
- The four-way interaction:

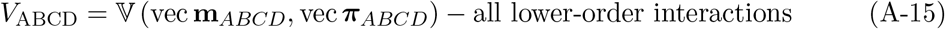

## Footnotes

1 In the R programming language, the `as.vector` and `as.matrix` commands implement some of this functionality.

2 That is obviously impossible, but the thought experiment is useful.

3 In one study that partitioned lifespan disparity using both measures, the results were very similar [11].

